# The Clinical Effectiveness of One-Dose HPV Vaccine: A meta-analysis of 902,368 vaccinated women

**DOI:** 10.1101/2023.08.17.23294214

**Authors:** Didik Setiawan, Nunuk Aries Nurulita, Sudewi Mukaromah Khoirunnisa, Maarten J Postma

## Abstract

**Introduction:** Although the effectiveness of the HPV vaccine has been comprehensively described, adherence to dosing and limited budget is one of the causes of delay in HPV vaccination implementation in a country. A one-dose HPV vaccine could possibly solve those issues since several studies show promising results.

**Areas covered:** This is a systematic review and meta-analysis focusing on the effectiveness of the one-dose HPV vaccine compared to two- and three doses of vaccination. We focused on clinical effectiveness, including prevention of HPV16, HPV18, and hrHPV infection, HSIL or ASC-H incidence, and CIN II/III incidence.

**Expert opinion:** Our review showed that a one-dose HPV vaccine could potentially be as effective as two-or three doses since it offers immunogenic protection up to 8 years follow up and also prevention on infection and pre-cancers incidence. However, more studies and an extended duration of existing studies are required in order to provide robust evidence of this recommendation.

## Introduction

Cervical cancer has becomes the second most common cancer globally and is mainly caused by Human Papillomavirus (HPV) infection^1^. This infection has been recognized to be responsible for about a quarter-million death-related to cervical cancer every year. Among the known HPV types. Some of HPV types (e.g. type 16 and 18) are categorized as carcinogenic HPV or high-risk HPV (hrHPV) while other types of HPV (e.g. type 6 and 11) are categorized as non-carcinogenic types or low-risk HPV (lrHPV)^2^. Therefore, prevention of hrHPV infection will, as a matter of course, protect against cervical cancer.

Since the nature of cervical cancer development has almost been understood perfectly^3^, the prevention strategies of cervical cancer have been introduced in several countries, considering HPV vaccination and cervical screening as primary and secondary prevention, respectively^4^. Previous studies showed that HPV vaccines are considerably effective in stimulating the antibody-specific hrHPV and potentially prevent cervical cancer in the future^5^.

Three commercial HPV vaccines are currently available in the market, each vaccine offers not only protection to HPV16 and HPV18 infection, as the leading cause of cervical cancer, but also some additional clinical benefits^6–8^. While quadrivalent vaccine also offers protection for HPV6 and HPV11 as the leading cause of genital warts, bivalent and nonavalent vaccines also provide additional protection to other hrHPV including HPV31, HPV33, HPV45, HPV52, and HPV58 as the additional cause of cervical cancer with different degrees of protection to each type of virus^9^.

During its initial introduction, HPV vaccines were suggested for three doses of administration to achieve some degree of protection against HPV infection and cervical cancer new cases^10^. During its application, also confirmed with continuing clinical trials, the immunogenicity profile of two doses of HPV vaccine was accidentally comparable with three doses of application^11^. These findings suggested a substantial reduction in cost-related vaccination since it has become one of the leading implementation barriers in several countries. The latest scattered findings from various studies showed that one dose of administration could potentially provide comparable results with two or even three doses of administration.

Several studies showed that the implementation of HPV vaccination as a cervical cancer prevention policy faces several issues such as coverage, acceptance, and the high price of the vaccine^12^. The reduction of cost-related vaccines will substantially influence the decision regarding implementing the policy in a country. Therefore, this study will initially start with a systematic review and meta-analysis that mainly focus on extracting information regarding the immunogenicity of the HPV vaccine that has been administered, both intentionally or unintentionally, for one dose compared to two or three doses of administration.

## Methods

We systematically searched for all studies that administered one dose of HPV vaccine intentionally or unintentionally from two electronic databases (PubMed and EMBASE). Two keywords (‘HPV vaccine’ AND ‘one dose’) and their respective MeSH term and text word were used to identify all related studies in the PubMed database. These similar keywords were also used in EMBASE database using exp (explosion search), ab (abstract), and ti (article title) commands. The last searching process was done on November 18^th^, 2022. in order to expand the search result, a snowball search was done by identifying the reference of both included studies and existing review articles. For risk of bias (RoB) assessment, Risk of Bias assessment tool version 2 and ROBINS-1 assessment tool for RCTs and pbservational studies, consecutively.

### Data collection and analysis

Since clinical outcomes are substantial in clinical decision-making, we only included studies that provide measurable clinical data. The results in this study was focused on the prevention of infection (both all hrHPV or HPV16 and HPV18 only) and prevention of pre-cancer/cancer development (HSIL or ASC-H and CIN II/III). Another inclusion criterion for this systematic review and meta-analysis is that the article is written in English. Two reviewers (DS and NAN) screened and assessed the detected studies independently, and any disagreement was discussed and solved with a third reviewer (MJP). Based on the Preferred Reporting Items for Systematic Reviews and Meta-Analyses (PRISMA) statement, we will extract the following data: authors, year published, country, study design, vaccine types, study population, number of subjects in each group, follow up time, outcomes and main finding of the included studies.

### Meta-analysis

To perform a meta-analysis, we extracted the incidence of HPV infection and pre-cervical cancers from both intervention groups (1 dose and the combination of two and or three doses). The HPV infection is differentiated into the infection caused by (1) HPV16 and HPV18 only and (2) all types of high-risk HPV (hrHPV). Moreover, other clinical outcomes in this meta-analysis have investigated the impact of one-dose HPV vaccine compared to 2 or 3 doses on (1) HSIL or ASC-H and (2) CIN II/III.

Risk Ratio (RR) was calculated from the number of events (infections and pre-cancers) in one-dose HPV vaccine groups compared to the groups who received two and or three doses of HPV vaccines using random-effects models to obtain the vaccine efficacy and or effectiveness. To deal with heterogeneity that could cause by the differences in sample characteristics and its methods, we estimated a heterogeneity test by calculating the I^2^ score according to Cochrane Q test results. This method presents a value of heterogeneity ranging from 0% to 100%. According to Cochrane’s recommendation, I^2^ of 50% or higher is considered substantial heterogeneity. This meta-analysis will be performed using RevMan 5.3.

## Results

### Screening Flow

From each database, this study found 405 and 494 from PubMed and Embase, respectively. After duplication removal (253 articles), there were 566 articles were excluded during abstract and title screening with the following reasons: studies are not about a single dose HPV vaccines (475 articles), communication articles including commentaries and letter to the editors (N=18), articles are not written in English (N=9), review articles (N=47), qualitative and case report article (N=3), official report documents (N=5), and clinical guideline and recommendation (N=9). During the full text screening, there were 64 out of 80 articles were excluded due to various reasons and 7 articles were included during snowball searching (Figure 1). Finally, 23 articles were included in the qualitative review and finally, there were only 11 articles were included in the meta-analysis since they provide explicit quantitative data on HPV vaccine effectiveness.

**Figure 1.**
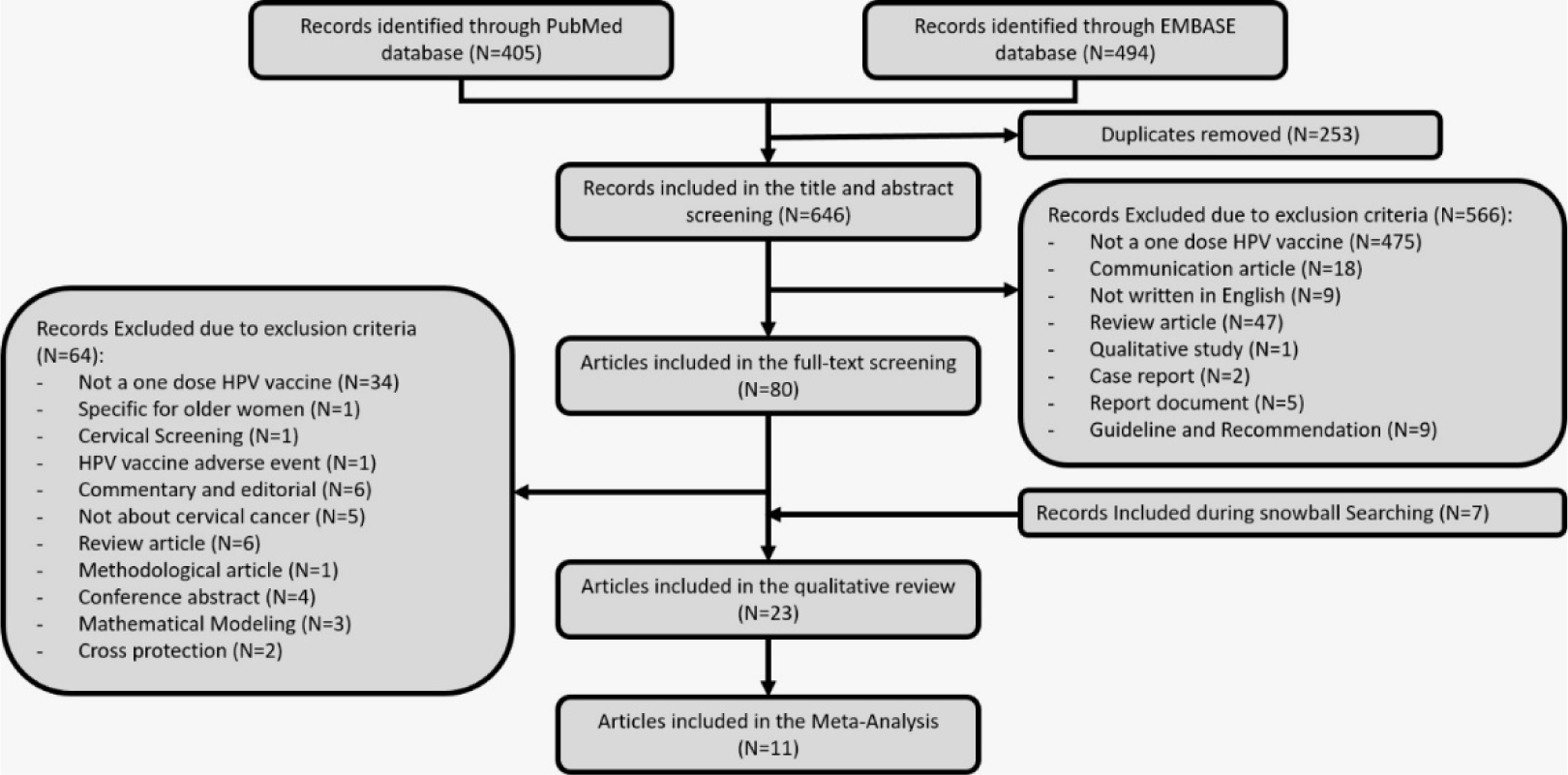
Screening Process

### Risk of Bias Assessment

The result of quality assessment for RCTs and observational studies is shown in Figure 2 and 3. The risk of bias assessment using RoB 2 showed majorities of the studies(1–5) presented a sufficient description of the randomization process. Meanwhile, three trials showed a high risk of bias due to deviations from the intended interventions. The outcomes of all the trials were estimated sufficiently and led the low bias by missing outcomes data. Five studies(1–4,6) depicted that the method of measuring the outcome was not inappropriate, and the outcome measurement did not differ between intervention groups. Some concerns in the selection of the reported results was shown in two studies(1,5). The overall result of the quality assessment showed that the four studies(1,5,7) had some concern of the risk of bias, and the remaining studies(2–4,6) demonstrated a high risk of bias.

**Figure 2.**
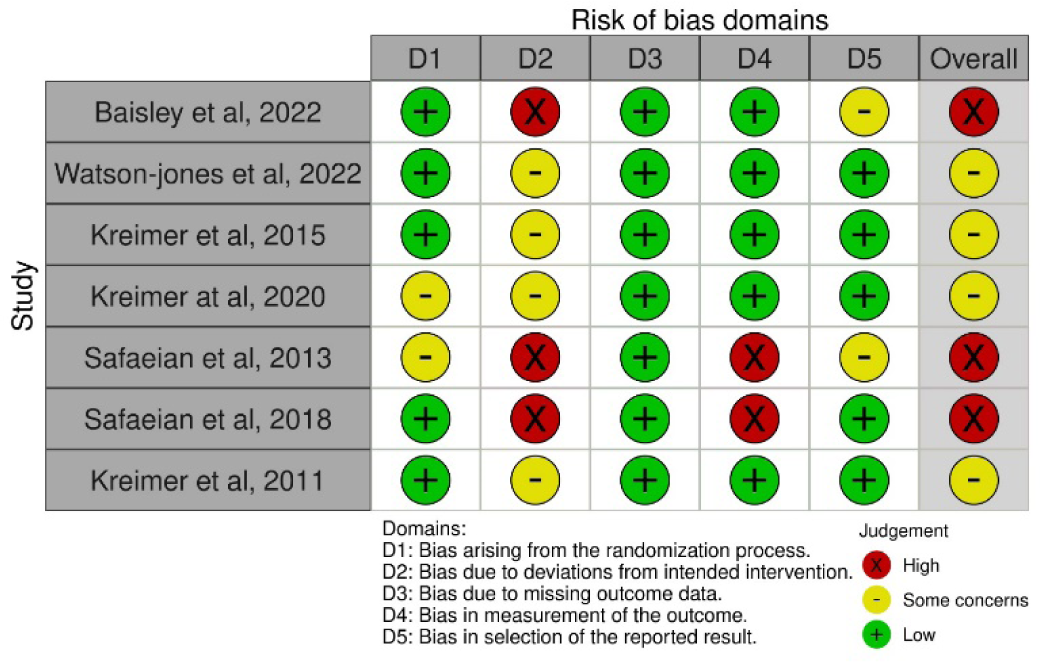
Risk of bias using RoB (Risk of Bias) version 2 for RCTs studies

**Figure 3.**
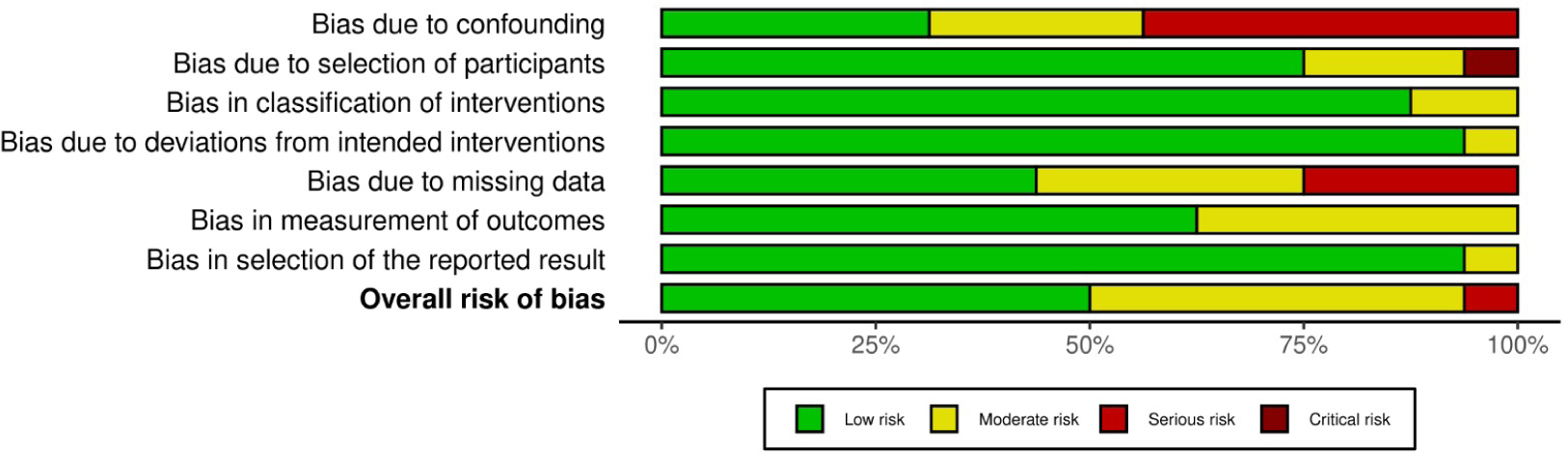
Risk of bias assessment using ROBINS-I for observational studies

The assessment results using ROBINS-i showed that there is a low risk of bias in the studies by presenting adequate reporting. However, some studies(8–14) had a serious concern on the bias due to the confounding. Most of the studies exhibited sufficient reporting on the selection of participants, classification of interventions, and low of risk of bias due to deviations from intended interventions. A high selection bias that arises due to exclusion of individuals with missing information about intervention status was found in four studies (10,13,15,16). Finally, the measurement of outcomes and selection of the reported result were sufficiently reported.

### Study Characteristics

The included studies were performed from various countries, including Australia^13–16^, India^17–19^, Canada^20^, Fiji^21^, United States^22,23^, Denmark^24^, Costa Rica^25–28^, Scotland^29,30^, Uganda^31^, Worldwide^32^, the Netherlands^33^ and Tanzania^34,35^. Two updated studies, one from India^18^ and another one from Costa Rica^26^, were included in the review but not in Meta-Analysis due to double counting concerns. Most included studies using observational cohort (11 studies)^13,15,30,36,16–19,21,23,24,29^ and Randomized Control Clinical Trials (RCTs) (4 studies)^26–28,32,34,35^. Furthermore, both Quadrivalent^13,14,24,36,15–21,23^ and Bivalent^25–33^ vaccines were almost equally studied. In addition, study comparing exclusively Bivalent versus Nonavalent^35^ and Bivalent, Quadrivalent and Nonavalent^34^ were also included in this review. There is a considerably wide variety of study population age (9 to 29 years old), the investigated group arms (unvaccinated to a complete three doses), duration of follow up (1 to 11 years long). Additionally, various clinical outcomes, both intermediate and endpoint, were investigated, including immunogenicity, infection prevention, and the prevention of pre-cancer and cancer itself.

Among 23 included studies, more than half of them conclusively mention that one-dose HPV vaccine provides similar vaccine effectiveness compared to two or three-doses of vaccination^17,18,32,34,35,37,19–21,24,25,27,29,31^. In addition, several studies suggest that antibody is available to protect the women who received one-dose HPV vaccine for at least 4 to 8 years, and immune memory was induced in that particular group^19,38–40^. However, three studies mentioned that the one-dose HPV vaccine did not significantly reduce the incidence of both pre-cancer and infection rates compared to unvaccinated group^13,14,29^. Two studies conclude that although one-dose HPV vaccine is immunogenic and reduces the incidence of pre-cancers, however, the protection of this particular dose is not equal to two or three doses ^15,33^.

**Table 1.** the characteristic of included studies.

| No | Author, Year | Location | Study design | vaccine types | Study population | Number of population on each group | Study period | Outcomes | Main findings |
| --- | --- | --- | --- | --- | --- | --- | --- | --- | --- |
| 1 | Gertig, 2013 | Australia | Retrospective cohort study using linked data from registries | QV | 12-13 y.o | Unvaccinated - 15192;<br>1 dose = 2568;<br>2 doses = 3412;<br>3 doses = 21199 | 5 years | Cervical Abnormalities (Histological and Cytological) | there was no significant reduction among those partially vaccinated (1 or 2 doses) compared to unvaccinated group |
| 2 | Crowe, 2014 | Australia | Case Control Analysis | QV | 11-27 y.o | Unvaccinated - 53761;<br>1 dose = 9649;<br>2 doses = 10950;<br>3 doses = 23106 | 3 years | CIN 2 or 3/ adenocarcinoma–in–situ (AIS)/ cancer | there was no statistically significance effectiveness of one dose HPV vaccine for high grade cervical abnormalities compared to unvaccinated group |
| 3 | Brotherton, 2015 | Australia | retrospective cohort study | QV | 26 or younger | 1 dose = 20,659;<br>2 doses = 27,500;<br>3 doses = 108,264 | 2.89 years (average) | CIN2, CIN3, AIS or mixed CIN3/AIS | Any number of doses (1, 2 or 3) was found to be associated with lower rates of high grade and low grade cytology diagnoses |
| 4 | Sankaranarayanan, 2015 | India | cohort study | QV | 10-18 y.o. | 1 dose = 4950;<br>2 doses (1 and 60) = 3452;<br>2 doses (1 and 180) = 4979;<br>3 doses = 4348 | 60 months | Immunogenicity, incidence and prevalence | The short- term protection afforded by one dose of HPV vaccine against persistent infection with HPV 16, 18, 6, and 11 is similar to that afforded by two or three doses of vaccine and merits further assessment. |
| 5 | Kim, 2016 | Canada | Nested Case-Control | QV | 10-11 y.o. | Unvaccinated - 5712;<br>1 dose = 327;<br>2 doses = 490;<br>3 doses = 3675 | 8 years | Abnormal cytology | The adjusted odds of high-grade cervical abnormalities with at least 1 dose of vaccination compared with no vaccination were lower |
| 6 | Toh, 2016 | Fiji | Prospective cohort study | QV | 15-19 y.o. | Unvaccinated = 32;<br>1 dose = 40;<br>2 doses = 59;<br>3 doses = 66 | 6 years | Seropositivity | A single dose of 4vHPV elicits antibodies that persisted for at least 6 years, and induced immune memory, suggesting possible protection against HPV vaccine types after a single dose of 4vHPV |
| 7 | Sankaranarayanan, 2018 | India | cohort study | QV | 10-18 y.o. | 1 dose = 4950;<br>2 doses (1 , 60 days) = 3452;<br>2 doses (1, 180 days) = 4979;<br>3 doses = 4348 | 7 years | Immunogenicity, incidence and prevalence of HPV infection | in our observational cohort study, one dose HPV vaccine is immunogenic and provides protection against HPV 16 and 18 infections similar to the two- and three-dose vaccine schedules |

### The effectiveness of HPV vaccine against HPV16 and HPV18 infection

Currently, most of the evidence shows that the prevalence of HPV16 and HPV18 infection in cervical cancer patients is the highest among other serotypes of HPV. Therefore, HPV16 and HPV18 infection incidence is closely related to cervical cancer cases. Six included studies describe that the risk of HPV16 and HPV18 infection among the one-dose group is slightly higher than the more-doses group (RR=1.55, CI95% 1.18-2.04, *p value* 0.002) (figure 4). this result is influenced mainly by a study from India^19^ and Scotland^30^ (weight 29.3% and 28.2%) where the study results were possibly underestimated the potential effectiveness among younger girls. furthermore, the comparison between one- and two doses of HPV vaccine also shows a differences in HPV16 and HPV18 infection rate between two groups (RR=1.18, CI95% 0.92-1.44, *p-value* 0.10) (Figure 5).

**Figure 4.**
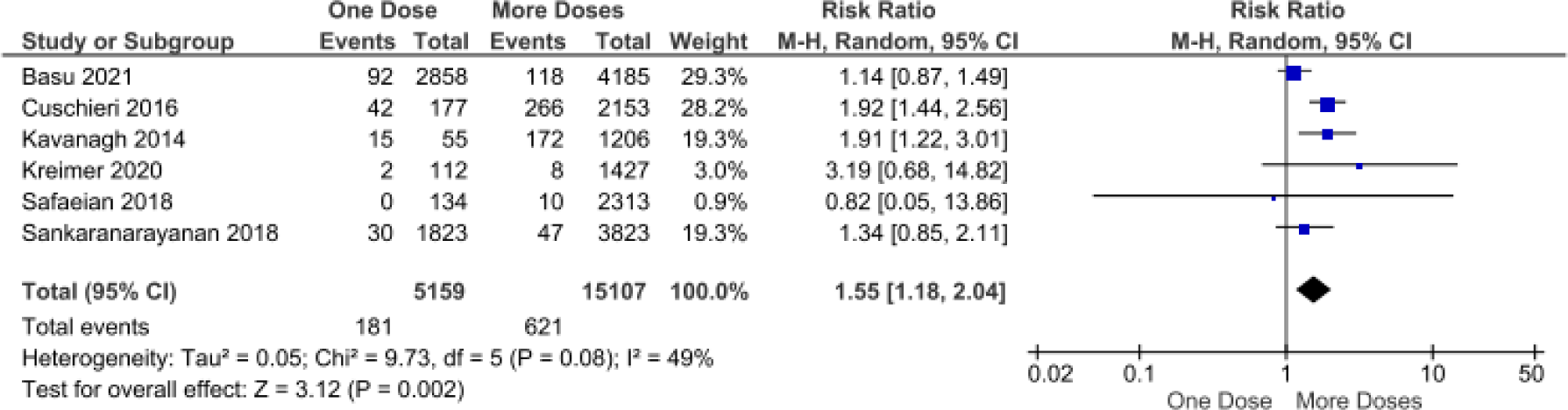
The effectiveness of one- and more-doses HPV vaccine on preventing HPV16 and HPV18 infection

**Figure 5.**
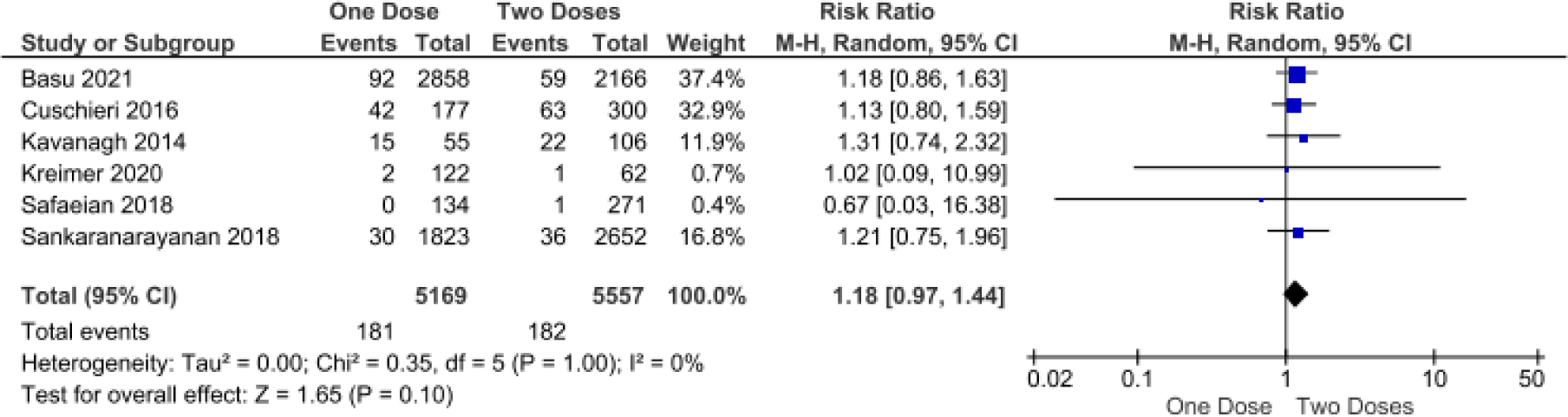
The effectiveness of one- and two-doses HPV vaccine on preventing HPV16 and HPV18 infection

### The effectiveness of HPV vaccine against hrHPV infection

In addition to HPV16 and HPV18, there are several types of HPV that have been known as high-risk HPV, including HPV31, HPV33, HPV45, HPV52, and HPV58. Those other types of HPV are included as hrHPV since they are also highly detected on cervical cancer patients ^41,42^. This study also explores the vaccine effectiveness in preventing the infection of hrHPV among vaccinated girls. This meta-analysis shows that the effectiveness of one-dose HPV vaccine on preventing hrHPV infection are slightly lower than more-doses (RR=1.27, CI95% 1.02-1.57, *p value* 0.03) (Figure 6) and two doses alone (RR=1.14, CI95% 1.03-1.27, *p value* 0.01) (Figure 7).

**Figure 6.**
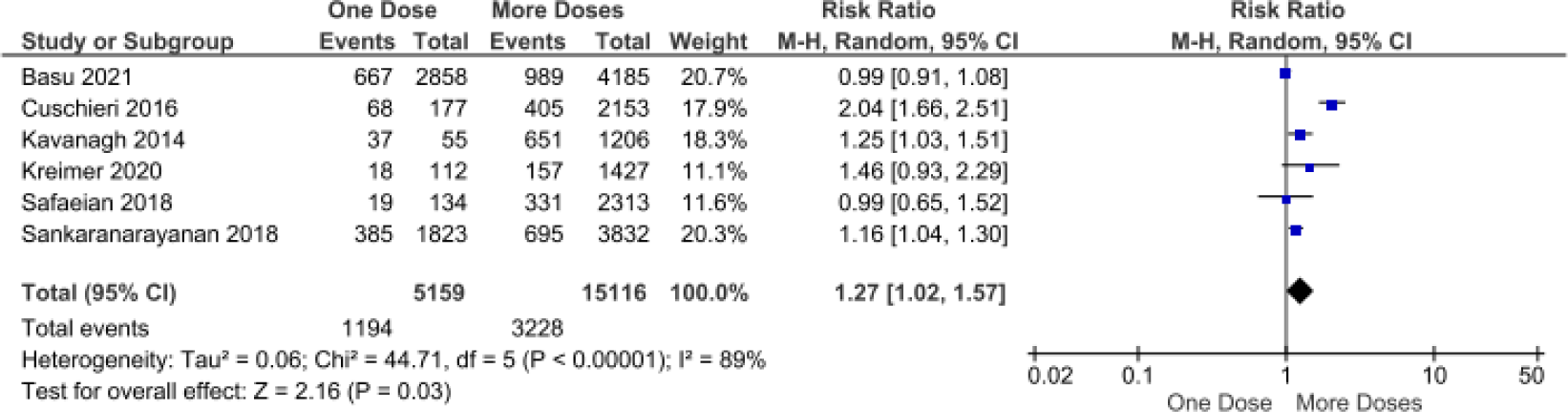
The effectiveness of one- and more-doses HPV vaccine on preventing hrHPV infection

**Figure 7.**
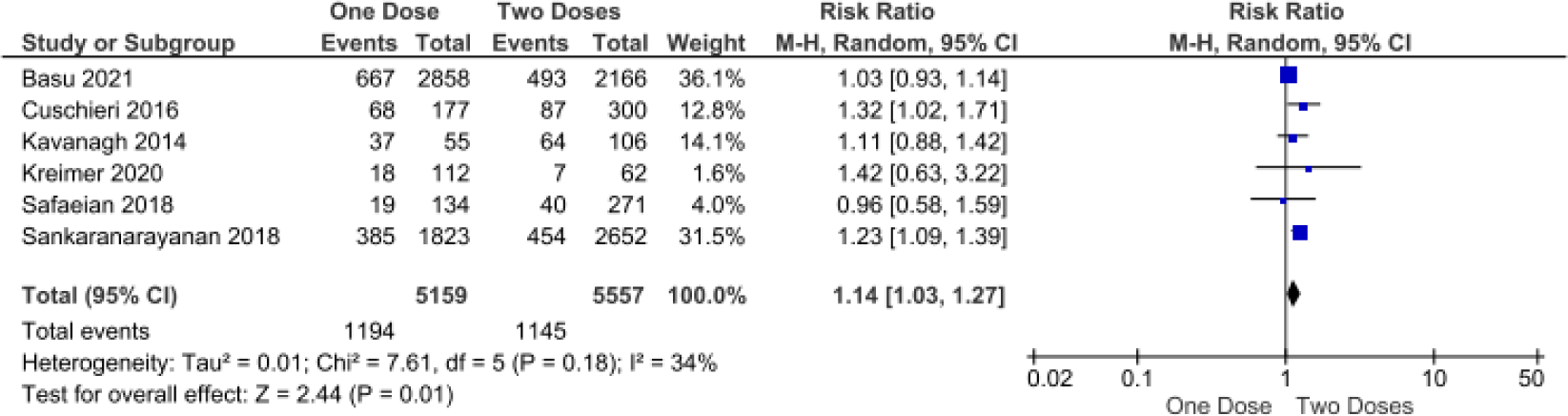
The effectiveness of one- and two-doses HPV vaccine on preventing hrHPV infection

### The effectiveness of HPV vaccine on preventing HSIL or ASC-H incidence

In the pap smear test, although Atypical Squamous Cell-H (ASC-H) is not included as a cancer cell, it could be part of High-Grade Squamous Intraepithelial Lesion (HSIL) which is included in a pre-cancer category. Therefore, a further confirmation test is required since HSIL may become cervical cancer if not treated quickly. In this study, the impact of one-dose HPV vaccine on preventing the incidence HSIL or ASC-H is apparently similar with more-doses (RR 1.29; 95% CI 0.88 – 1.88; *p-value* 0.19) (Figure 8) or two-doses alone (RR 1.01; 95% CI 0.74 – 1.37; *p-value* 0.97) (Figure 9). Although this result was only generated from a few studies, the individual studies showed almost comparable results and patterns, especially comparing one dose versus more doses. However, these analyses found heterogeneity among supported studies (I^2^ of 88% and 51%).

**Figure 8.**
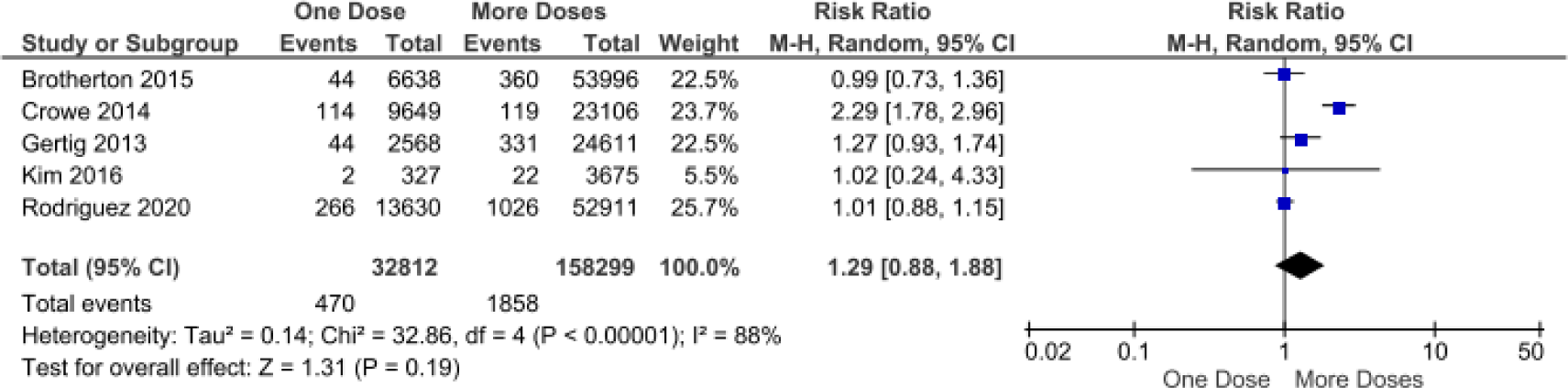
The effectiveness of one- and more-doses HPV vaccine on preventing HSIL or ASC-H incidence

**Figure 9.**
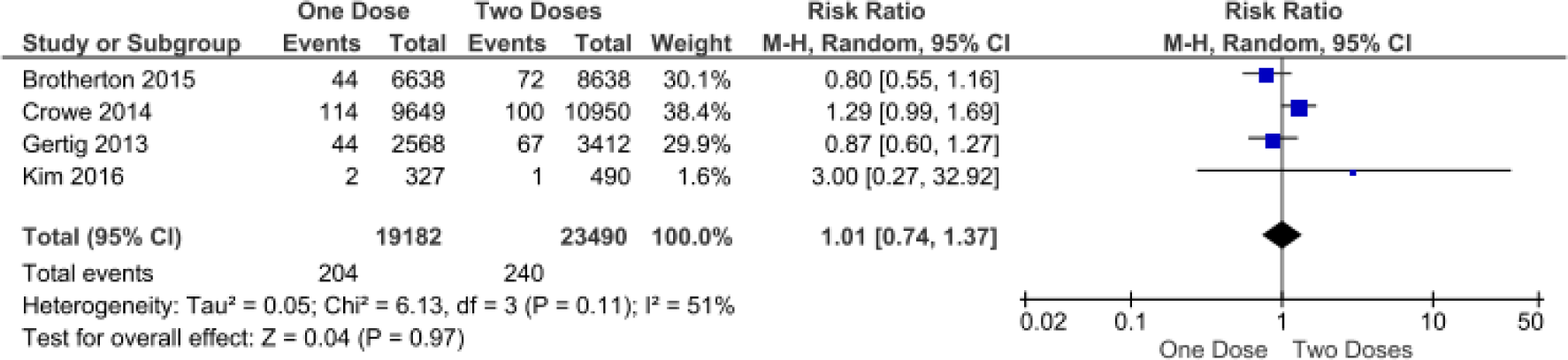
The effectiveness of one- and two-doses HPV vaccine on preventing HSIL or ASC-H incidence

### The effectiveness of HPV vaccine on preventing CIN II/III incidence

Cervical Intraepithelial Neoplasia (CIN) are precancerous stages where some abnormal cell grows on the surface of the cervix, and it can be found during cervical screening, for example, pap smear test and Visual Inspection using Acetic Acid (VIA). The classification of CIN is determined according to the proportion of the cervix, particularly the epithelial surface. CIN 2 usually describes that dysplasia affects about one-third to two-thirds of the epithelium. While CIN 3 represents that more than two-thirds of the epithelial layer of the cervix is changed and it is considered the most severe form of CIN. This meta-analysis finds that women who received one-dose HPV vaccine showed comparable effectiveness with women who received more doses to prevent CIN II/III (RR=1.54, CI95% 0.91-2.62, *p value* 0.11). However, there is high heterogeneity among studies in this analysis (I^2^ of 94%) (Figure 10).

**Figure 10.**
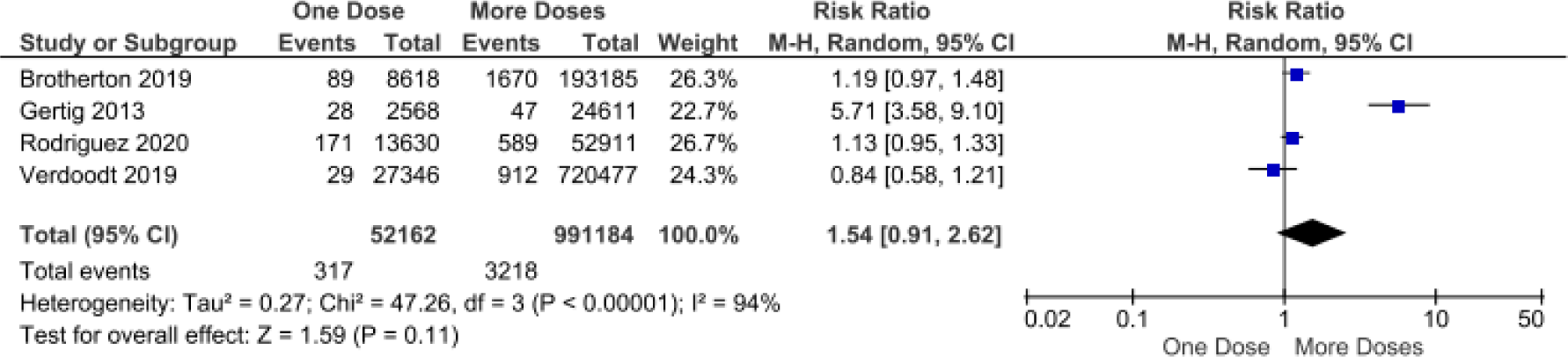
The effectiveness of one- and more-doses HPV vaccine on preventing CIN II/III incidence

## Discussion

The nationwide implementation of HPV vaccination in a country often faces problems, including the low adherence to second or third dose and limited budget to provide complete doses of the HPV vaccine, especially in low- and middle-income countries. A one-dose HPV vaccine could solve these issues significantly reduce those problems. However, valid and quantitative evidence on the efficacy or effectiveness of one-dose HPV vaccine should be available beforehand to help the decision-making process. This systematic review focus on the effectiveness of the one-dose HPV vaccine compared to more doses (two- and three-doses) on protecting infection and pre-cancer incidence. From this systematic review and meta-analysis, we found that most of the included studies support that one-dose HPV vaccine induces HPV-specific antibodies up to 8 years, providing immunogenic memory and as effective as two-or more doses of vaccination on preventing infection and pre-cancer condition. However, few studies explain that the antibodies are not equal to two-or three doses, and the reduction in infection rate and the incidence of pre-cancer is not significant. Therefore, more studies and more extended study periods are required to provide a definitive conclusion of the effectiveness of the one dose HPV vaccine.

This study strengthens the previous systematic review on one-dose HPV vaccine^43^ since this study includes not only Randomized Controlled clinical Trials but also observational studies that have a longer duration of follow-up. In addition, this review also considers the end-point clinical outcomes, including pre-cancer and cancer incidence, which are more critical in the perspective of patients, clinicians, and other decision-makers. Ultimately, this meta-analysis substantially provides a quantitative pooled analysis of relative risk between one-dose versus more doses and two-doses of HPV vaccines across available studies in the world. In addition, this review does not support the previous review from Markowitz et al., which clearly explained that the immunogenicity of the one-dose HPV vaccine is inferior to three doses.

This study extracted pooled quantitative vaccine effectiveness data on preventing both infection and pre cancers incidence. One of the exciting findings in this meta-analysis is that the effectiveness of a one-dose HPV vaccine is generally comparable with more doses in pre-cancer prevention but not on infection prevention. Several studies, mainly from Scotland and India, do not support the hypothesis of similar effectiveness between one-versus more-doses HPV vaccines on preventing hrHPV infection. Both studies explain that one-dose HPV vaccine does not sufficiently induce cross-protection, particularly on HPV31, HPV 33, and HPV 45. These studies could be the main drive on why one-dose HPV vaccine does not as effective as more doses on preventing hrHPV infection.

Since several countries have implemented a two-dose HPV vaccination schedule, this review also evaluates the head-to-head comparison between one-versus two-dose HPV vaccines. According to our meta-analysis, the one-dose HPV vaccine provides comparable effectiveness with the two-dose HPV vaccine on the prevention of not only HPV16 and HPV18 but also HSIL or ASC-H incidence. On the other hand, although comparable effectiveness between two alternatives could not be presented in terms of hrHPV infection prevention, two studies that did not support this particular comparison explain that one-dose HPV vaccine has comparable effectiveness with more doses (the combination of two-and three-doses). This phenomenon could cause by the limited number of the one-dose group or the number of incidences in the one-group arm.

One of the limitations of this study is that there were only 23 included studies and several of them are updated studies. Moreover, only 4-6 studies can be included in each meta-analysis. This condition restricts our study when sub-group analysis should be performed to evaluate the impact of one dose on a specific age group. Furthermore, this study did not assess the effectiveness of the one-dose HPV vaccine on the endpoint clinical outcome, which is cervical cancer incidence. However, although longer follow-up time is required before deciding on implementing a one-dose HPV vaccine schedule is made, these consistent findings provide a promising comparable benefits between one- and more-doses of HPV vaccines.

## Data Availability

All relevant data are within the manuscript and its Supporting Information files.

